# Datasheet for the IDHea Primary Eye Care Dataset: A Real-World Ocular Imaging Resource for Research

**DOI:** 10.1101/2025.04.29.25326706

**Authors:** Reena Chopra, Anya Guzman, Yi Sing Hsiao, Christopher Lee, Julia Jolly, Juho Uotila, Nessa Pantfoerder, Ian Douglas, Anthony P. Khawaja, Pearse A. Keane, Matthew P. Lungren, Julia Coelho, Timothy Bossie, Mary K. Durbin

## Abstract

**Purpose:** Real-world ocular imaging datasets are essential for advancing research in artificial intelligence (AI), autonomous disease screening, and clinical decision support. The Primary Eye Care dataset is a large-scale collection of de-identified retinal imaging data from routine optometric care, made available through the Institute for Digital Health (IDHea)—a secure research platform established by Topcon Healthcare, Inc. This dataset provides an opportunity to study eye health in a community setting and will be available via this cloud-based platform.

**Methods:** Data were collected and de-identified from individuals who underwent imaging as part of their routine care across 40 optometry practices in the United States and one practice in Australia. The dataset includes threedimensional optical coherence tomography (OCT), and color fundus photographs acquired using Maestro devices (Topcon Corp., Tokyo, Japan), along with demographic data including age and sex. Imaging data were converted to DICOM format, and OCT analysis metrics such as retinal layer thicknesses were derived. Additional labels including image quality, vessel metrics, and retinal pigment score were generated using open-source AI models.

**Results:** TThe dataset comprises 873,291 image acquisitions from 276,061 subjects with a mean age of 43.8 years (standard deviation = 19.5). 48.7% were female, 36.2% as male, and 15.1% not reported. Most OCT scans followed the 12 x 9 mm 3D Wide protocol (86.3%), with additional 3D Macula, 3D Disc, anterior segment, radial, and line scans. 59,049 subjects (21.4%) had two or more scans separated by ***≥***365 days. Pre-processed metrics and AI-derived labels, such as TopQ image quality scores, glaucoma risk score, and AutoMorph features are included. 89.4% of OCT scans scored above 25 on the TopQ scale, indicating reliable image quality. A propensity score-matched test subset (~10%) was held out to enable consistent benchmarking across studies.

**Conclusion:** The Primary Eye Care Dataset provides a large-scale, real-world collection of ocular imaging data, reflecting a largely healthy, community-based population attending routine optometric visits. This makes it particularly valuable for developing AI models aimed at early detection and prevention at the population level, where most eyes are healthy, and disease prevalence is low. Data access is governed by an independent committee to ensure ethical and responsible use; more information is available at IDHea.net

## Introduction

Real-world data on normal ocular anatomy and function play a crucial role in advancing our understanding of eye health and disease. By capturing the normal variations and physiological characteristics of eyes in routine screening settings, baseline references can be established that are essential for identifying deviations indicative of pathology [1]. This type of data is invaluable for developing and validating diagnostic tools, particularly those based on artificial intelligence (AI) which target screening settings, as it provides a comprehensive foundation for training algorithms to recognize a spectrum of normal anatomy, hence optimizing specificity. Furthermore, real-world data from routine clinical settings could offer insights into the prevalence and progression of eye conditions across diverse populations, enabling more accurate and personalized approaches to eye care. The availability of such data is fundamental to improving early detection, screening programs, and ultimately, patient outcomes.

Although medical imaging is increasingly recognized as a valuable research asset, several challenges must be addressed to create comprehensive and accessible repositories of such data. These include navigating data ownership complexities that limit direct access to these data sources, implementing HIPAA-compliant de-identification protocols, developing robust data cleaning and curation pipelines to ensure dataset quality, and establishing secure research platforms and governance processes to ensure patient privacy. After these issues have been addressed, the data needs to be formatted to well-adopted community standards and data models, such as Digital Imaging and Communications in Medicine (DICOM), in structured databases to enable large-scale data sharing in a research-ready format.

The Primary Eye Care dataset addresses key challenges in accessing real-world ocular imaging data and represents a significant advancement in the field. It includes an extensive collection of optical coherence tomography (OCT) scans and color fundus photographs (CFP), captured during routine visits to optometry practices in the United States and Australia. With a diverse population, the dataset offers a comprehensive snapshot of ocular health in community-based settings.

This dataset offers the potential to capture both normal anatomical variation and incidental or early signs of pathology, making it a valuable resource for a broad range of research. Applications include phenotyping healthy eyes, identifying early disease biomarkers, studying the prevalence and progression of ocular conditions, and developing or validating AI models for detection and diagnosis. The inclusion of demographic information further enables population-level analyses of how factors such as age, sex, and regional variation influence eye health.

Recognizing the potential of this real-world dataset, Topcon Healthcare, Inc. (THI) has established the Institute for Digital Health (IDHea) to de-identify, curate, and securely host the dataset for research access. IDHea encompasses a data repository and secure workspace dedicated to research and innovation using ocular imaging and diagnostic data. The primary aim of IDHea is to transform population screening data into a research-ready format. The key attributes of IDHea are:

- **Fast**: Streamlining data access requests to provide rapid access to a large repository of data to accelerate discovery and innovation.
- **Accessible**: Providing an intuitive and user-friendly platform that supports a diverse range of research initiatives, from exploratory analyses to machine learning model development, that can be accessed from anywhere, in the cloud.
- **Safe**: Employing industry-standard security protocols and infrastructure to provide the highest level of privacy and ensure compliance with health data protection regulations.
- **Transparent**: Maintaining a clear, open, and publicly available governance framework to build trust and ensure fair access to data. All data access requests are reviewed by an independent committee.

IDHea is designed to support fast, efficient, collaborative research by offering seamless access to enriched, well-organized datasets. Researchers across academic and industrial sectors can request access through a transparent governance process overseen by an independent Data Access and Governance (DAG) committee. The DAG is responsible for reviewing and approving data access requests, ensuring that data usage aligns with ethical guidelines and the broader goal of advancing ocular research to improve patient outcomes.

This datasheet provides an overview of the IDHea Primary Eye Care dataset, outlining the collection, composition, and characteristics, and follows guidelines set out by Rostamzadeh et al. 2022 [2].

## Datasheet

### Dataset composition

The Primary Eye Care dataset is composed of routinely collected data in optometry practices across 11 states in the US, and a single optometry practice in Victoria, Australia. At present, thirteen sites are included in this dataset, where each site encompasses one or more practice locations within the same geographical area, typically under the care of a single lead optometrist. In total, the dataset covers 41 individual practice locations. All images were acquired using 3D Optical Coherence Tomography 3D OCT-1 (Types: Maestro or Maestro2) (Topcon Corp., Tokyo, Japan) devices, comprising paired OCT and color photographs (Figure 1). The dataset is designed to reflect real-world clinical practice, capturing a wide range of demographics and regional diversity. Table 1 provides an overview of the locations included in the dataset and the demographic characteristics.

**Table 1.** Characteristics of sites included in the Primary Eye Care dataset. Age and sex are reported at a scan-level. NR: Not reported.

| Site | Locations | State | Acquisition date range | Unique subjects | Unique images | Age (mean, SD) | Sex (reported) |
| --- | --- | --- | --- | --- | --- | --- | --- |
| AUS | 1 | Victoria, Australia | 2017–2021 | 4,463 | 25,744 | 65.8 (15.6) | Female: 0.0%<br>Male: 0.0%<br>NR: 100.0% |
| Site001 | 1 | New York | 2016–February 2025 | 7,664 | 44,227 | 38.2 (11.1) | Female: 59.3%<br>Male: 40.3%<br>NR: 0.4% |
| Site003 | 2 | Wisconsin | 2012–August 2024 | 6,572 | 33,787 | 44.0 (18.7) | Female: 53.4%<br>Male: 45.9%<br>NR: 0.7% |
| Site004 | 2 | Georgia | 2021–March 2025 | 11,419 | 28,919 | 44.7 (15.7) | Female: 62.1%<br>Male: 37.3%<br>NR: 0.6% |
| Site005 | 2 | New Jersey | 2020–February 2025 | 10,725 | 52,735 | 49.3 (19.4) | Female: 52.3%<br>Male: 40.3%<br>NR: 7.4% |
| Site006 | 2 | Texas | 2019–August 2024 | 5,492 | 20,565 | 54.3 (21.9) | Female: 57.9%<br>Male: 41.2%<br>NR: 0.9% |
| Site007 | 1 | Florida | 2020–2022 | 1,456 | 4,775 | 58.6 (17.3) | Female: 53.5%<br>Male: 40.1%<br>NR: 6.4% |
| Site008 | 1 | New Jersey | 2012–February 2025 | 4,240 | 21,259 | 42.4 (17.7) | Female: 54.6%<br>Male: 44.3%<br>NR: 1.2% |
| Site009 | 9 | New Jersey | 2019–December 2024 | 86,283 | 254,235 | 39.4 (18.9) | Female: 53.9%<br>Male: 36.0%<br>NR: 10.1% |
| Site010 | 4 | North Carolina | 2020–February 2025 | 4,842 | 10,915 | 47.9 (15.8) | Female: 47.7%<br>Male: 51.3%<br>NR: 1.0% |
| Site011 | 8 | Massachusetts, Rhode Island, New Hampshire | 2019–February 2025 | 25,487 | 76,009 | 49.6 (18.0) | Female: 53.1%<br>Male: 41.1%<br>NR: 5.7% |
| Site012 | 1 | Tennessee | 2020–February 2025 | 6,352 | 29,762 | 44.7 (20.9) | Female: 57.2%<br>Male: 40.0%<br>NR: 2.8% |
| Site013 | 7 | New York | 2017–2023 | 101,066 | 270,359 | 42.6 (19.9) | Female: 42.6%<br>Male: 34.0%<br>NR: 23.4% |
| **Total** | 41 | – | – | **276,061** | **873,291** | **43.8 (19.5)** | **Female: 49.3%**<br>**Male: 36.3%**<br>**NR: 14.3%** |

**Fig. 1.**
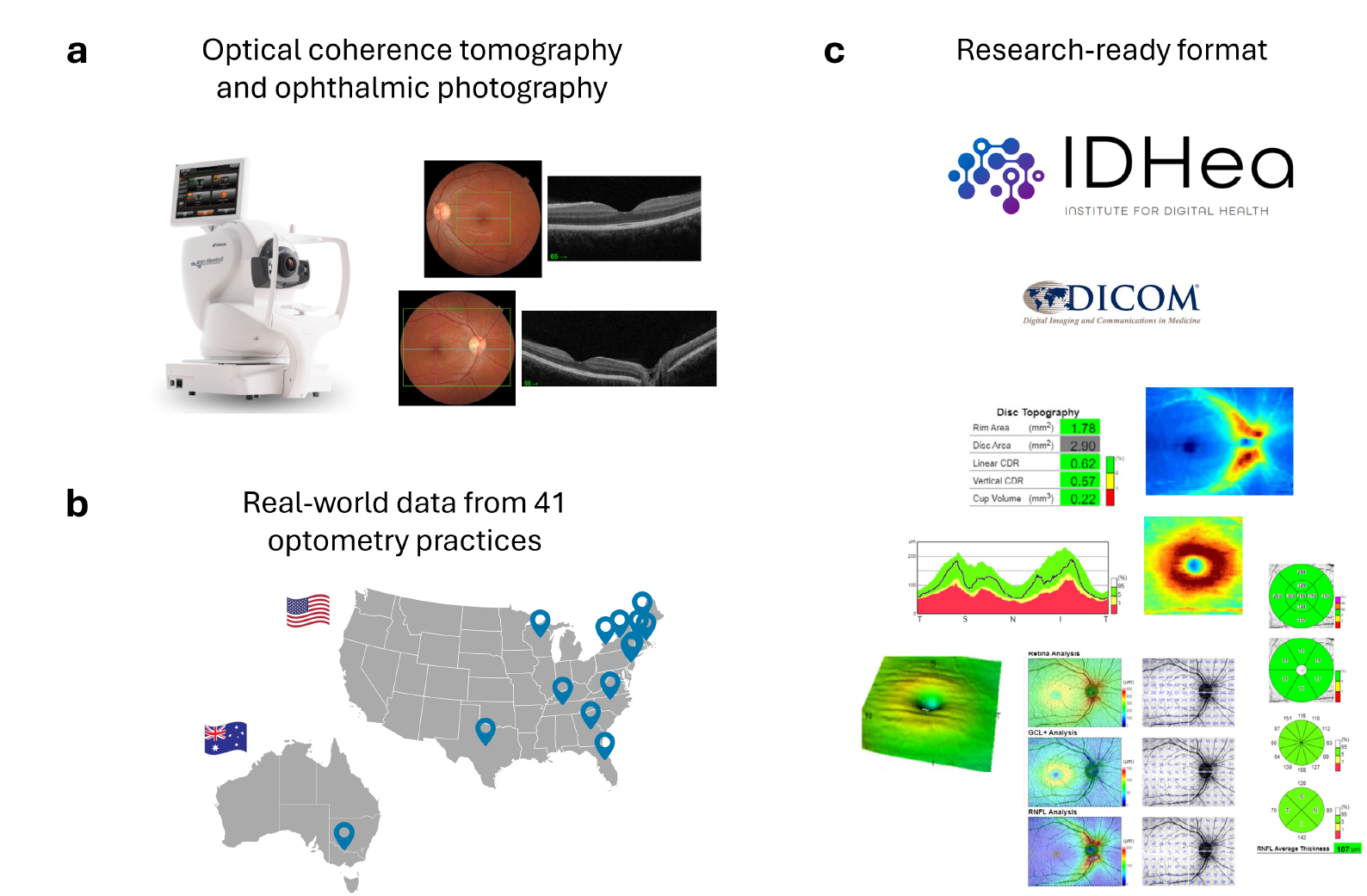
Development of a research-ready ocular imaging dataset for IDHea. a) OCT and color fundus photographs were acquired as part of routine clinical workflows. b) Data were collected from geographically diverse optometry practices across the United States and Australia. c) Imaging data were curated into a research-ready format through DICOM conversion, extraction of OCT metrics, and generation of AI-derived features. The structured dataset is made available via the IDHea platform for research and model development.

### Ethics

Data were collected, de-identified, and curated by THI. Data sharing agreements were established between THI and each participating optometry practice to outline the terms under which de-identified data can be utilized for research while safeguarding subject confidentiality and ensuring compliance with ethical and regulatory standards.

Additionally, approval was obtained from the Advarra Institutional Review Board (IRB) (Columbia, MD, USA) for the retrospective collection of de-identified data in the US (IRB # CR00611155), which requires annual renewal. IRB approval was also obtained for the Australia dataset from the Bellberry Human Research Ethics Committee (Protocol # 2022-04-345-FR-1).

### Data collection and de-identification

The data were collected retrospectively from routine optometry visits. Imaging was offered as part of standard optometric care as a paid service, with some sites providing imaging free of charge for certain age groups. The dataset includes both retinal and anterior segment OCTs, as well as color photographs. OCTs were captured using various scan modes, including: 3D (horizontal or vertical) scan mode with a scan resolution of 512 pixels width and 128 B-scans, Radial scan mode (12 B-scans spaced 30^*?*^ apart, 5-Line Cross with each line having 1024 pixels width, or a single Line scan of 1024 pixels width. For 3D scans, Fixation was either Wide (12 x 9 mm scans), Disc (6 x 6 mm scans), Macula (6 x 6 mm or 7 x 7 mm scans), or External (6 x 6 mm or 12 x 9 mm). Radial, 5-Line Cross, and Line scan modes were either a Macula, Disc, or External fixation and had scan widths of of 3 mm, 6 mm, or 9 mm. Anterior segment OCTs were captured using External fixation and were accompanied by anterior eye photographs.

Prior to any research use, the data were de-identified in accordance with the Health Insurance Portability and Accountability Act (HIPAA) Privacy Rule’s Safe Harbor method (45 C.F.R. § 164.514(b)(2)), which specifies the removal of 18 direct and quasi-identifiers to render data non-identifiable. Under HIPAA, once data have been deidentified according to this standard, they are no longer considered protected health information (PHI) and may be used or shared for research purposes without patient authorization.

The de-identification process was performed using a standardized process. Patient identifiers were first combined with a site-or location-specific secret salt, a random value added to each identifier before hashing to enhance security and prevent re-identification through precomputed hash lookups. For Site003, Site006, Site010, and Site012, a single salt was used across all locations within each site, as patients may attend multiple locations and are assigned the same patient identifier throughout. The salted identifiers were then processed using a one-way cryptographic hash function (SHA-256) and truncated to a fixed length for consistency and compatibility. The resulting text representation of the hash replaced the original identifier and is thus known as the subject identifier. Within image files, subject identifiers were base32 encoded and truncated to 18 characters, while filenames were shortened to 8 characters after base32 encoding. Dates of birth were removed and replaced with a default date of 01/01/1910, with image capture dates adjusted relative to this reference. This approach maintains the chronological order of longitudinal scans and preserves the ability to calculate the subject’s age to the nearest month and the time interval between the scans. Ages above 90 years are recorded as 90 to reduce the risk of re-identification.

Some eyes have multiple image acquisitions captured during the same visit. To preserve the temporal sequence of captures, the exact time of image acquisition has been retained within the metadata, allowing researchers to reconstruct the order of imaging events within a visit.

### Pre-processing and labeling

To ensure data quality, images intended for device testing or demonstration purposes were excluded. A data-cleaning process was implemented to maintain metadata consistency, addressing cases where a subject’s name, date of birth, or sex had been corrected or changed during subsequent visits. This process ensured that all prior images were updated to align with the most recent meta-data. This cleaning procedure was successfully implemented for 29 of 41 locations. Due to limitations in the available database records, it could not be applied to all locations in this dataset version, specifically Australia, Site007, Site013, two locations at Site009 and one location at Site011.

To standardize imaging formats, all images were converted to DICOM format. This conversion ensures compatibility with industry standards, enhancing interoperability and facilitating integration across research platforms and analytical tools. Ophthalmic Photography 8-Bit Images and Ophthalmic Tomography Images are stored in JPEG2000 Transfer Syntax to avoid data loss in conversion. Relevant DICOM attributes include pixel spacing, slice thickness, and additional metadata supporting quantitative imaging analysis. For 3D OCT volumes, retinal layer contour coordinates enabling layer and multi-layer thickness calculations as well as the generation of two-dimensional enface projections and retinal thickness maps, are stored in Service-Object-Pair (SOP) Instances in accordance with the Height Map Segmentation Information Object Definition (IOD). The datasets carry consistent cross-references between files, and each object is conformant with IODs as described in DICOM PS3.3 release 2025a [3–5].

To reduce the computational burden on researchers, OCT images have been pre-processed using Topcon’s OCT analysis software, which automatically generates a comprehensive set of retinal thickness measurements for specific layers—retinal nerve fiber layer (RNFL), ganglion cell layer (GCL) and inner plexiform layer denoted as GCL+, the sum of the RNFL and GCL+, denoted as GCL++, and the total retina—measured across spatial grids including the Early Treatment Diabetic Retinopathy Study (ETDRS) grid, the macula 6-sector grid, the Super Pixel grid, and the circumpapillary circle (3.4mm diameter circle centered using the disc center position) [6, 7]. This eliminates the need for researchers to run their own segmentation algorithms and standardizes down-stream analysis. Other retinal thickness measurements can be derived, using DICOM files containing segmentation information, between any of several segmented layers, including the internal limiting membrane (ILM), RNFL/GCL boundary, GCL, inner plexiform layer/inner nuclear layer (IPL/INL) boundary, outer plexiform layer (OPL), external limiting membrane (ELM), photoreceptor inner segment/outer segment (IS/OS) junction, outer segment/retinal pigment epithelium (OS/RPE) boundary, and Bruch’s membrane (BM).

Additionally, outputs from several AI models are available to facilitate research without requiring users to implement their own image analysis pipelines. AutoMorph, an AI-based model for CFP, is included to assess image quality and extract features related to the optic disc and blood vessels [8]. Retinal pigment score (RPS) was applied to CFP deemed good or usable quality by AutoMorph [9]. The dataset also includes the Multi-factorial OCT Score (MOS), a validated risk score ranging from 0 to 100 that estimates the likelihood of glaucoma, based on a Japanese study and validated in a US population [10]. Finally, we provide an estimate of axial length developed at Topcon, which utilizes acquisition parameters such as mirror position and retinal location within the image, which may serve as a useful proxy for identifying short and long eyes in the absence of refractive error measurements [11]. By including these pre-processed outputs, we aim to streamline research efforts, allowing users to focus on higher-level analyses without the need for additional computational processing.

To facilitate AI model validation, a standardized and representative test subset was held out using propensity score matching (PSM) [12], enabling consistent bench-marking and comparison of algorithm performance across studies. This subset, comprising approximately 10% of individuals from each optometry practice, was selected to reflect the broader population in terms of key covariates: age, sex, laterality, and scan protocol. PSM was applied to balance these characteristics, ensuring that the selected test set is representative of the full dataset while maintaining the real-world diversity of routine optometric imaging. Access to this subset is limited and requires submission of a formal request through the same governance and review process as is used to request access to the full dataset.

### Dataset characteristics

Summary statistics are reported prior to separating the test subset. Overall, the dataset comprises 873,291 image acquisitions from 276,061 individuals. Among the included patients, the mean age was 43.8 years (SD = 19.5) (Figure 2), with 48.7% reported as female, 36.2% as male, and 15.1% not reported. A total of 62,974 (22.8%), 59,049 (21.4%), and 34,609 (12.5%) subjects had two or more scans separated by at least 183, 365, and 730 days, respectively.

**Fig. 2.**
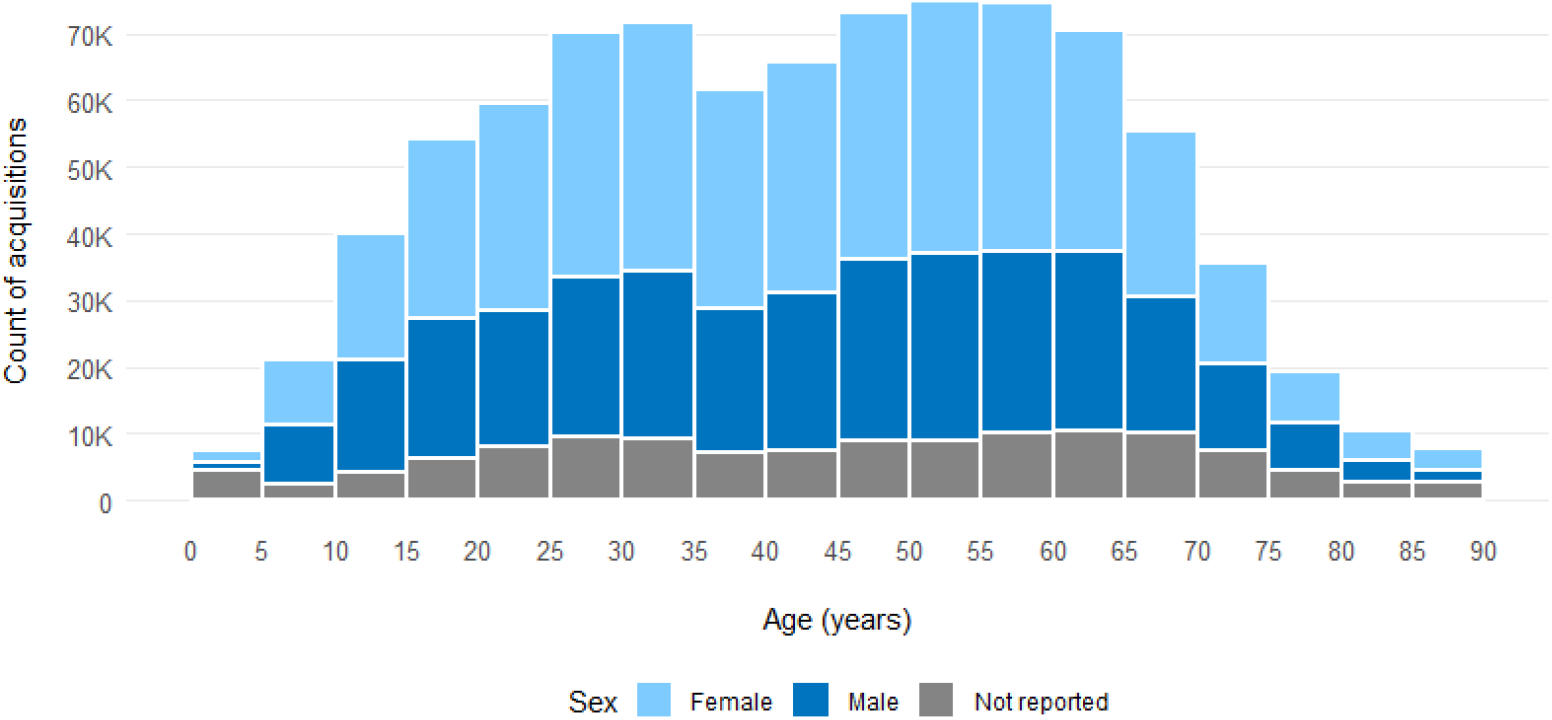
Age distribution, stacked by sex, all sites combined.

The dataset includes 753,396 (86.3%) 3D Wide (12 × 9 mm) scans, 25,093 (2.9%) 3D Macula (6 × 6 mm or 7 x 7mm) scans, and 29,509 3D Disc (3.4%) (6 × 6 mm) scans. 868,421 (99.4%) acquisitions included a paired OCT and color photo, 4,854 (0.6%) contained an OCT only, and 16 included a color photo only. 808,164 (92.5%) scans were 3D volumes, 44,521 (5.1%) were 5-Line Cross scans, 11,555 (1.3%) were Radial scans, and 9,035 (1.0%) were single Line scans. 20,058 (2.3%) scans were anterior segment OCTs and included all scan modes.

OCT-derived metrics include a range of retinal layer thickness measurements, such as ETDRS central subfield retinal thickness: mean = 240.2 µm, SD = 39.5 µm), average circumpapillary RNFL thickness (mean = 99.9 µm, SD = 21.7 µm), and average GCL+ thickness across the macula 6 sectors (mean = 70.2 µm, SD = 10.5 µm). Axial length estimates ranged from 15.9 to 34.2 mm, with a mean (SD) of 24.6 (1.3) mm. This mean is slightly higher than population averages reported in the literature [13], likely reflecting the optometric setting in which the data were collected, which may be skewed toward individuals with myopia and thus longer axial lengths. TopQ image quality assessment, a unitless measure which corresponds to the retinal and inner choroidal image quality [14], was applied to all OCT images and had a mean (SD) score of 40.0 (13.5) with 89.4% scoring above 25, indicating reliable quality scans suitable for advanced analysis across most scans in the dataset (Figure 3). Threshold values specific to individual scan types are provided in Table 2.

**Table 2.** Scan reliability by scan type based on TopQ image quality thresholds. Scans were categorized as “reliable” or “not reliable” based on predefined TopQ image quality thresholds specific to each scan type. A total of 801,420 scans had a scan type and TopQ score available for applying these thresholds. No cutoff value is applied to the other scans which are not described above. 3D(H) represents scans with horizontal lines scans.

| Scan Type (TopQ Threshold) | Not reliable, n (%) | Reliable, n (%) |
| --- | --- | --- |
| 3D(H) Wide ($\geq 25$) | 85,008 (11.3) | 665,137 (88.7) |
| 3D Macula ($\geq 28$) | 676 (3.1) | 21,090 (96.9) |
| 3D Disc ($\geq 30$) | 1,478 (5.0) | 28,031 (95.0) |

**Fig. 3.**
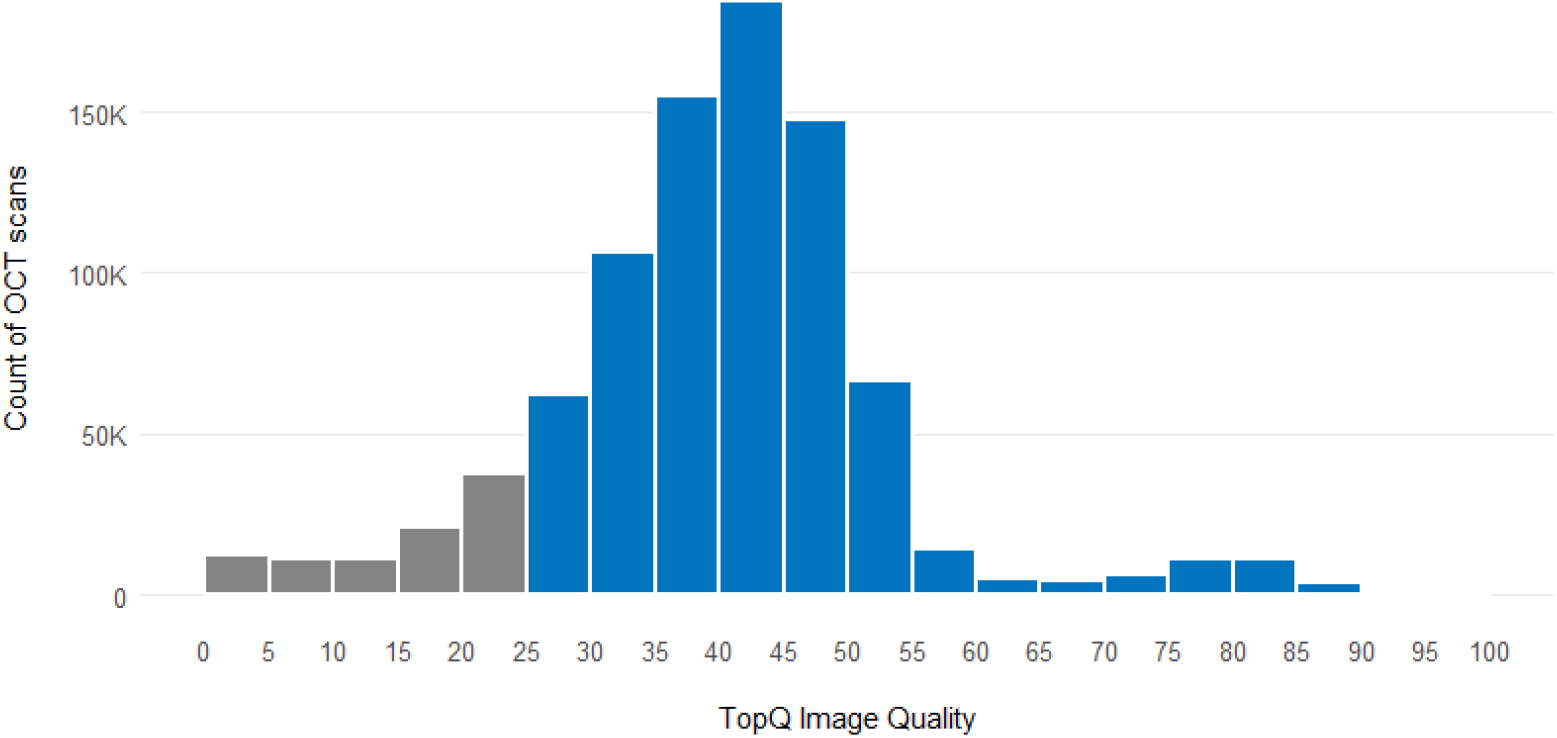
TopQ image quality distribution. A TopQ quality of 25 and above is considered to be of reliable quality for advanced analysis for the majority of scans.

The MOS was applied to all 3D Wide OCT scans. To remove segmentation artifacts and mispositioned images, the following filtering criteria was also applied, including: age *≥* 40 years; sufficient image quality (TopQ *≥* 18); disc and fovea positions within reasonable bounds of the image (2.4–9.6 mm width, 1.8–7.2 mm height); disc area measurements *≤* 6.5 mm^2^; and complete peri-papillary RNFL thickness measurements without missing or zero values in any sector. 38,625 (4.4%) scans from 25,016 (9.1%) subjects achieved a score of *≥* 90, indicating a potential risk of glaucomatous damage.

AutoMorph, an algorithm designed for retinal image quality assessment, was applied to all color photographs, including anterior segment photos, categorizing 56.4% as “good” quality, 15.5% as “usable”, and 28.1% as “bad” quality.

### Dataset versioning

This datasheet summarizes the first version of the dataset release, comprising data collected up to March 2025. There are plans to release new major versions of the dataset periodically with additional data collected from new sites and/or existing sites. For existing sites, new records will be matched to previously collected data using consistent de-identification methods, ensuring that each unique subject retains the same identifier across versions. Sub-versions will be released on an ongoing basis which will include additions such as labels to an existing version.

### Data access and governance

The IDHea Primary Eye Care dataset is available to researchers across academic and industrial sectors through a structured application process designed to ensure ethical, secure, and responsible data use. Applicants must submit their research proposal outlining their objectives, methodologies, and anticipated outcomes for evaluation.

The IDHea Data Access and Governance committee, an independent body, is responsible for reviewing data access requests and ensuring alignment with ethical principles and governance policies. Comprising experts in clinical practice, research, and technology, the committee provides multidisciplinary oversight to safeguard responsible data use. All members serve in a personal capacity, acting independently of their institutional or commercial affiliations to ensure impartial and unbiased decision-making.

The committee establishes criteria for evaluating research and development applications, focusing on adherence to IDHea’s governance policies, ethical standards, and alignment with previously approved or precedent use cases. Each application undergoes a structured review to confirm eligibility, including verification of the applicant’s identity and affiliation and the suitability of the proposed use. Similar or identical use cases may be approved for multiple applicants, and assessments are grounded in policy and precedent rather than scientific evaluation. This process ensures consistency, fairness, and responsible use of the dataset.

Transparency is a fundamental principle of the committee’s operations. Every data access request is reviewed against predefined governance policies, with documented findings ensuring clear accountability. Where applications are approved and data access is granted, the project title and name of the lead researcher will be published online at IDHea.net to uphold integrity and trust in the governance process and research use.

A tiered access model with discounts for academic users supports broad participation in research while ensuring the sustainability of the data infrastructure. Access to additional resources, such as expert annotations or AI model-generated labels, will incur fees for both academic and commercial users, reflecting the added cost and effort involved in producing and maintaining these enriched datasets.

### Secure cloud computing environment

The dataset is hosted within a secure workspace on Microsoft Azure Databricks [15], ensuring a controlled environment with no data egress while providing integrated tools for data analysis, machine learning, and collaborative development. Microsoft Azure Databricks is HITRUST CSF certified and is designed to support compliance with standards like HIPAA, GDPR, and prevailing industry standards [16]. Researchers benefit from interactive notebooks, SQL-based querying, custom dashboards, and scalable compute clusters, all within a unified platform designed to streamline large-scale data science (Figure 4). While raw data remains within the IDHea secure workspace, users can export their trained model weights enabling them to apply their models in external workflows. To further protect data integrity, agreements are in place with researchers to ensure that any exports do not include elements that would allow for reconstruction or replication of the original dataset.

**Fig. 4.**
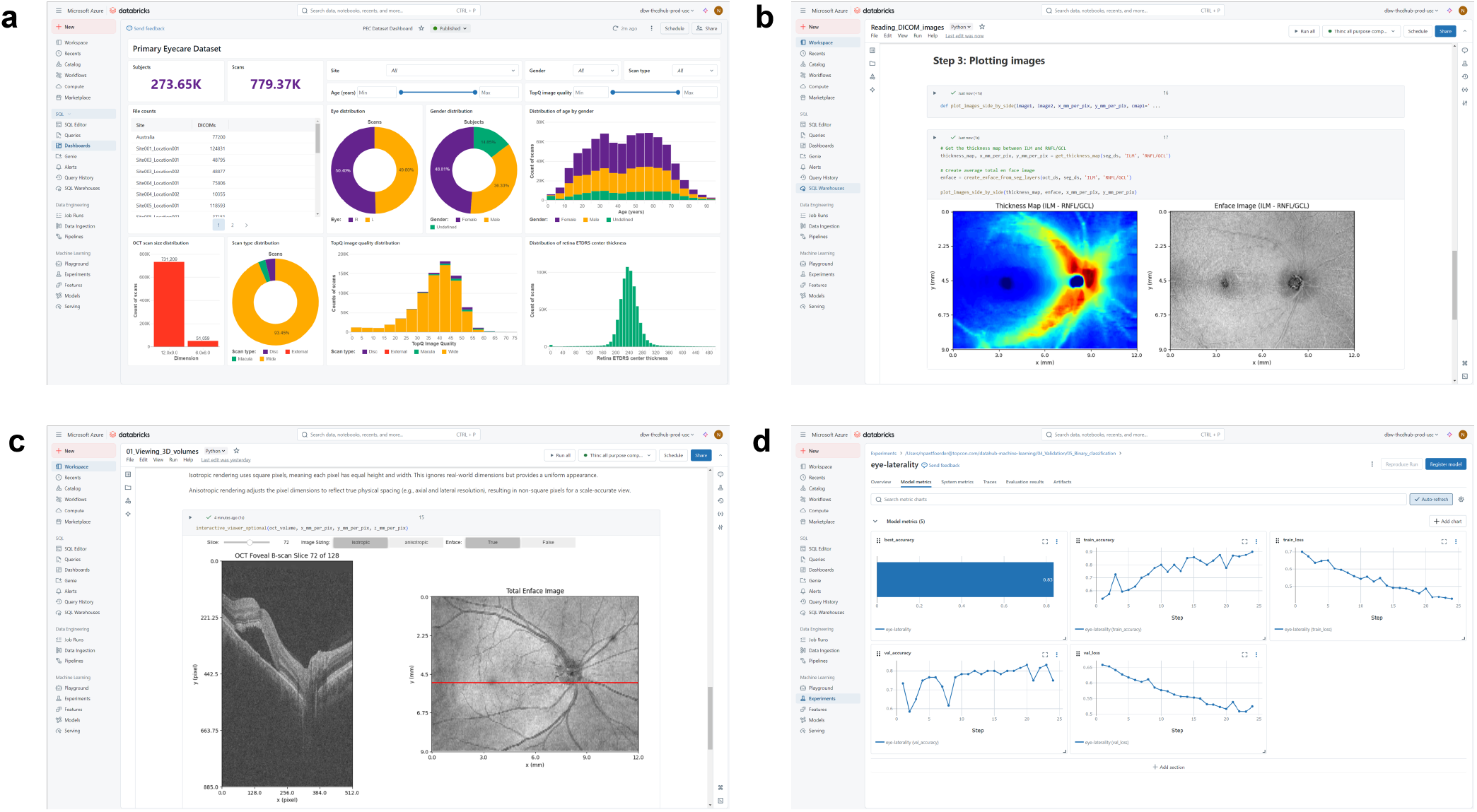
Microsoft Azure Databricks workspace. a) Interactive dashboards for data exploration. b) Reading segmentation DICOM files to generate enface projections and retinal thickness maps. c) Scrolling through OCT volumes using interactive elements coded using Python in Notebooks. d) Tracking the performance of machine learning experiments during training and validation.

To streamline research workflows, example notebooks are provided within the secure workspace, demonstrating how to read files, pre-process images, and perform basic machine learning development and evaluation. These notebooks offer step-by-step guidance for researchers, facilitating efficient data exploration and model development without requiring extensive setup.

Additionally, a cloud-based computing infrastructure enables access to GPUs for high-performance computing, supporting advanced computational tasks and democratizing access to powerful resources. In addition, cost management tools are available, enabling researchers to monitor and optimize compute usage for sustainable, cost-effective access to resources.

### Strengths and limitations

A key strength of this dataset is its large-scale, real-world representation of geographically diverse populations, providing valuable insights into routine optometric imaging. The inclusion of paired OCT and color fundus photographs, along with demographic data, enables multimodal analyses and population-level insights into ocular health. The dataset’s longitudinal structure, with many individuals having follow-up visits, supports studies exploring trends over time leading to better understanding of disease progression and opportunities for early intervention.

All images were acquired using Topcon Maestro devices and converted to DICOM format, which ensures compatibility with clinical imaging standards and provides rich metadata for quantitative, reproducible analysis. The inclusion of pre-processed OCT segmentation metrics, AI-derived image quality scores and vessel metrics, and glaucoma risk assessments reduces the computational burden on researchers, facilitating immediate analysis and the ability to compare and even combine results. AutoMorph categorized 71.4% of color photographs as good or usable quality. The distribution of fundus image quality likely reflects clinical workflow factors, where the primary focus was on acquiring high-quality OCT scans rather than optimizing fundus photography. Consistent with this, OCT image quality was high overall, with 89.4% of scans achieving a TopQ score *≥* 25. Since OCT image quality is independent of CFP image quality, scans with poor AutoMorph scores may still be suitable for studies focused solely on OCT data. For studies involving CFP analysis, AutoMorph scores provide a useful guide to image reliability.

Researchers benefit from access to a secure, cloud-based workspace that combines scalable compute resources, interactive notebooks, and tools for collaborative development, making it easier to conduct high-performance analysis without extensive infrastructure. To support reproducibility and fair comparison of AI models, the dataset includes a propensity score–matched test subset, representing approximately 10% of individuals. This shared subset enables consistent benchmarking across studies, helping to standardize model evaluation. Furthermore, regular dataset updates are planned to enhance its scope and utility.

Access is governed through a transparent oversight process, supporting both academic and commercial use while ensuring responsible data stewardship. The dataset will continue to expand through periodic updates, incorporating new data and enriched labels to further strengthen its value for the research community.

Current limitations include the absence of diagnostic labels, though future versions aim to incorporate structured diagnostic data from linked electronic health records and independent expert grading. In addition, a portion of the dataset contains missing demographic data, particularly for sex, and lacks structured race or ethnicity data, limiting some subgroup analyses. Finally, although the dataset spans 11 U.S. states and one site in Australia, certain regions may be overrepresented, which could affect the generalizability of findings to broader populations.

## Summary

The IDHea Primary Eye Care Dataset provides a large-scale, real-world collection of ocular imaging and demographic data, serving as a valuable resource for research in AI-driven diagnostics and population eye health studies. Unlike hospital-based or case-control datasets, it reflects an unselected, community-based population— largely healthy individuals attending routine optometric visits—representing the setting where AI models for early disease detection and prevention are most urgently needed. The geographic diversity across multiple U.S. states and Australia further enhances its generalizability for population-level applications.

This dataset builds upon existing real-world ocular imaging resources that have significantly advanced largescale eye research. The Illinois Ophthalmic Database Atlas (I-ODA) offers a multi-modal, longitudinal dataset that enables detailed clinical studies and disease progression analysis [17]. INSIGHT, a UK-based health data research hub, aggregates anonymized ocular images from a range of vendors and includes OCT and CFP from Topcon devices to support AI-driven disease detection and healthcare innovation [18–20]. The UK Biobank provides extensive ocular imaging data, also including OCT and CFP from a Topcon device, linked to genetic and health records, allowing for large-scale population health studies [21]. The IDHea Primary Eye Care dataset complements these efforts by focusing on primary eye care imaging, capturing routine optometric visits and providing a valuable benchmark for AI model development and validation.

With transparent governance and secure cloud access, researchers can analyze data within a controlled workspace while exporting trained model weights for external application. Regular updates will further expand the dataset’s scope, enhancing its utility for AI development, epidemiological studies, and clinical research. By reflecting real-world primary eye care, this dataset supports the development of models and research grounded in typical imaging workflows and patient populations, increasing the likelihood that findings will generalize and translate effectively into future decision support systems, influence daily clinical practice, and ultimately improve patient outcomes.

Data access can be requested by submitting an application at IDHea.net.

## Acknowledgements

We thank the following optometrists and practices for their contributions to the dataset: Dr Justin Bazan, Dr Norman Brandon, Dr Fabius Clements, Dr Charles Cummins, Dr Brittany Degler, Dr Randi Frankl, Dr Ryan Fritsch, Dr Jason Hendrix, Dr Susan Johnson, Dr Will Ling, Dr Matthew Marsich, Dr Theresa Radtke, and Dr Bradford Ripps.

We also gratefully acknowledge the following individuals for their thoughtful feedback and review of the manuscript: Marco Miranda, Carole McCallum, Kerry Goetz, Jacqueline Armani, Ray Everett, and Yvonne Bentley.

## Declarations

### Data availability

Applications for data access can be submitted via IDHea.net A sample dataset is freely available for download following registration.

### Code availability

Example notebooks written in Python are available within the Databricks environment.

### Author contribution

R.C., C.L., J.J., J.C., T.B., M.K.D. conceptualized the project. R.C., A.G., Y.H., C.L., J.J., N.P., J.U., M.K.D. collected, curated and processed the data. R.C., A.G., Y.H. performed the analysis and visualization. R.C. drafted the manuscript. All authors reviewed and edited the manuscript. J.C. and M.K.D. supervised the project.

